# Forecasting Clinical Risk from Textual Time Series: Structuring Narratives for Temporal AI in Healthcare

**DOI:** 10.1101/2025.11.01.25339295

**Authors:** Shahriar Noroozizadeh, Sayantan Kumar, Jeremy C. Weiss

## Abstract

Clinical case reports encode temporal patient trajectories that are often underexploited by traditional machine learning methods relying on structured data. In this work, we introduce the forecasting problem from textual time series, where timestamped clinical findings–extracted via an LLM-assisted annotation pipeline–serve as the primary input for prediction. We systematically evaluate a diverse suite of models, including fine-tuned decoder-based large language models and encoder-based transformers, on tasks of event occurrence prediction, temporal ordering, and survival analysis. Our experiments reveal that encoder-based models consistently achieve higher F1 scores and superior temporal concordance for short- and long-horizon event forecasting, while fine-tuned masking approaches enhance ranking performance. In contrast, instruction-tuned decoder models demonstrate a relative advantage in survival analysis, especially in early prognosis settings. Our sensitivity analyses further demonstrate the importance of time ordering, which requires clinical time series construction, as compared to text ordering, the format of the text inputs that LLMs are classically trained on. This highlights the additional benefit that can be ascertained from time-ordered corpora, with implications for temporal tasks in the era of widespread LLM use.

## Introduction

Healthcare disparities persist globally, with preventable deaths from conditions like sepsis disproportionately affecting underserved populations who often present to underresourced facilities with limited access to specialized expertise. In these settings, much of the critical diagnostic and prognostic information exists only in unstructured clinical narratives—case reports, discharge summaries, and progress notes—as comprehensive structured data infrastructure may be unavailable (Anzalone et al. 2025; Seinen et al. 2025). While machine learning approaches have shown promise on structured data, with recent work demonstrating that incorporating text alongside structured inputs significantly improves predictive performance (Kline et al. 2022), large language models struggle with clinical risk estimation, especially in patient-facing scenarios (Wong et al. 2025). This gap highlights the need for automated risk forecasting from textual clinical records that could democratize access to expert-level clinical reasoning, particularly in resource-constrained environments where timely specialist consultation is not available. The challenge of LLM risk estimation motivates a structured treatment of risk forecasting and survival modeling from unstructured narrative sources.

Among textual sources, retrospective case reports serve as a rich training ground for developing temporally-aware AI systems that can eventually be deployed on real-time clinical notes. Case reports provide holistic accounts of clinical trajectories with explicit temporal reasoning—exactly the type of structured thinking needed for real-time clinical decision support. However, their narrative structure interleaves temporally unordered information, making them difficult to apply in forecasting tasks. Without identifying the time of occurrence for each event, models are prone to causal leakage—using information not available at the time of prediction. Solving this temporal reasoning challenge in case reports establishes the foundation for deployment in live clinical environments where such capabilities could improve patient outcomes.

One might ask whether large pretrained language models can already perform forecasting from clinical narratives, given their exposure to biomedical texts (Peng, Chen, and Lu 2020; Weber et al. 2024). However, these models have architectural limitations for temporal reasoning tasks. Encoder-based models apply random masking that rarely yields temporally coherent representations, while decoder-style models mask text in linear sequence, modeling events in text order rather than time order. Our experiments demonstrate these limitations quantitatively–for example, using the time-line in Figure 1, the probability of recovering a valid time-ordered mask is *<* 0.002%, and decoder models achieve low temporal concordance (e.g., *c* = 0.23), leading to suboptimal forecasting accuracy without task-specific restructuring.

**Figure 1.**
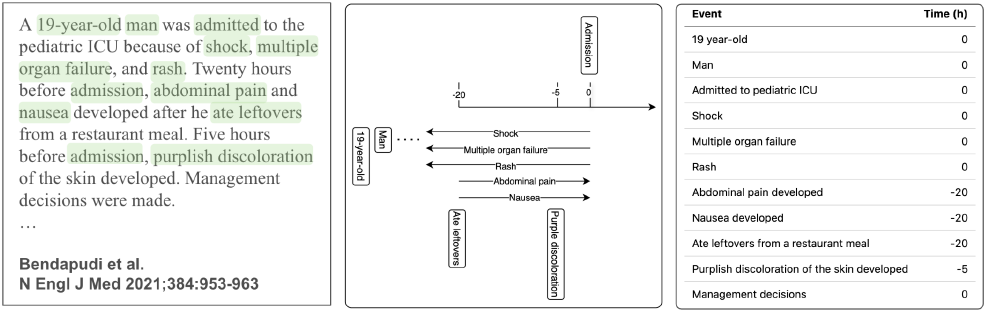
Example case report (left) with temporal representations as timelines (middle) and text-ordered, event-time tuples (right).

To address these temporal reasoning challenges, we develop a forecasting framework that transforms free-text narratives into textual time series—sequences of (event, time) tuples and makes explicit the temporal structure of patient trajectories. Building on recent annotation work (Wang and Weiss 2025), we propose a hybrid pipeline combining rule-based heuristics and LLM-assisted extraction to recover temporally localized clinical findings, validated on real clinical notes with expert verification.

Our work builds on extensive research in clinical NLP and temporal reasoning (Kohane 1987; Leeuwenberg and Moens 2020; Cheng and Weiss 2023), but shifts the emphasis from extraction to downstream clinical forecasting. While prior efforts have focused on entity recognition and temporal relation extraction (Zhou and Hripcsak 2007; Uzuner et al. 2011; Sun, Rumshisky, and Uzuner 2013), few have systematically assessed how such representations influence downstream tasks such as risk prediction and survival modeling. We bridge this gap by formalizing textual time series representations and evaluating their effectiveness for temporally grounded clinical prediction.

We empirically assess this framework across multiple tasks and modeling strategies, benchmarking encoder and decoder models—both fine-tuned and prompted versions—on three forecasting tasks: event prediction, survival analysis, and temporal ordering of future clinical events. Our results show no single model dominates across all tasks, highlighting the importance of aligning model architecture with forecasting objectives. We further examine how input ordering (time vs. narrative) impacts generalization and analyze the sensitivity of models to missing historical context through systematic masking experiments.

In summary, this paper makes the following contributions: (i) **Annotation and Extraction Pipeline:** Building on prior methods for extracting information from the PubMed Open Access corpus (Noroozizadeh and Weiss 2025), this work introduces a pipeline that transforms case reports into textual time series via regular expression filtering and LLM-assisted extraction, yielding temporally anchored (event, time) tuples while minimizing causal leakage by restricting input to past events; (ii) **Comprehensive Model Comparison:** We conduct a comprehensive evaluation of encoder- and instruction-tuned decoder-based models—using both finetuned MLP heads and prompting—across event prediction, temporal ordering, and survival analysis tasks. Results show no single model excels universally, underscoring the need to align model choice with the specific forecasting objective; (iii) **Temporal- versus Text- Ordering:** We investigate the impact of annotation order by comparing training on time-ordered versus text-ordered data. Results show that preserving the narrative’s natural order can improve generalization to external datasets, while time-based ordering can enhance ranking performance; (iv) **Sensitivity Analysis of Temporal Masking:** We conduct systematic dropout experiments by randomly masking parts of the clinical history to assess their impact on forecasting and event ordering. While higher masking levels reduce classification performance (F1), the concordance index remains largely stable, highlighting differing sensitivities of binary prediction and ranking tasks to historical context in textual time series; (v) **Methodological Framework for Temporal Clinical Forecasting:** Our approach demonstrates how narrative clinical texts can be systematically converted into structured temporal representations for forecasting tasks, providing a replicable methodology that could be adapted to other clinical and non-clinical text sources.

## Related Works

### Temporal Information Extraction from Clinical Text

Extracting timelines from clinical narratives is a challenging biomedical NLP task. The i2b2 2012 challenge introduced annotations datasets for temporal relation extraction from discharge summaries (Uzuner et al. 2011). Subsequent methods linked clinical events to timestamps or temporal expressions (Leeuwenberg and Moens 2020; Frattallone-Llado et al. 2024), typically using pre-defined event spans. We adopt the approach of Noroozizadeh and Weiss (2025) to assign timepoints to findings from full-length case reports, enabling finer temporal resolution. Unlike methods using structured EHR data, we focus solely on text—crucial for sources like PubMed that lack structured metadata. By directly supervising event-time alignment, we overcome limitations of span-based annotations (Rosenbloom et al. 2011). This aligns with growing emphasis on temporality in sepsis and critical care phenotyping (Johnson et al. 2018; Kamran et al. 2024), particularly given high missingness rates in structured data (Johnson et al. 2023; Seinen et al. 2025).

### Predictive Modeling with Clinical Text

Clinical text has been used for outcome prediction tasks like mortality and readmission, with models like ClinicalBERT achieving high levels of performance on EHR notes (Huang, Altosaar, and Ranganath 2019; Gu et al. 2021). Text captures complementary information—symptoms and social factors—not found in structured codes. However, traditional approaches using full-text or bag-of-words representations obscure temporal dynamics. Our approach preserves event order within narratives, treating data as time series. This aligns with recent efforts adapting LLMs for time series tasks, such as Time-LLM mapping numeric sequences to tokens (Jin et al. 2023). To our knowledge, this is the first work applying LLMs to text-derived time series in clinical domains. While prior sepsis prediction models rely on vitals or scores like SOFA and SAPS (Hou et al. 2020; Noroozizadeh, Weiss, and Chen 2023), our method is complementary—leveraging narrative descriptions that include clinician interpretations and context. This is valuable when case reports or patient histories are available but structured data is sparse or unavailable.

### Large Language Models in Healthcare

Recent LLMs such as *GPT-3* and *GPT-4* have enabled applications from medical QA to note summarization. These models encode substantial medical knowledge and reasoning capacity—*GPT-4* demonstrates strong performance on board exams and can support patient record abstraction for quality reporting (Boussina et al. 2024). However, their use in clinical forecasting remains largely unexplored. Our benchmark tests LLMs via zero- and few-shot prompting to assess clinical event forecasting from patient narratives, comparing results to fine-tuned models to quantify the gap between general-purpose and task-specific modeling. This contributes evidence that while LLMs hold broad medical knowledge, fine-tuning and structured inputs are often required for clinical reliability (Huang, Altosaar, and Ranganath 2019; Gu et al. 2021). Some studies suggest domain-specific tuning does not always improve over foundation models (Jeong et al. 2024), reinforcing the need for task-specific evaluations like ours.

## Methods^1^

We next describe our dataset construction and annotation pipeline, define the forecasting and survival prediction tasks, detail the modeling approaches evaluated, and present the sensitivity analyses used to assess robustness (Figure 2).

**Figure 2.**
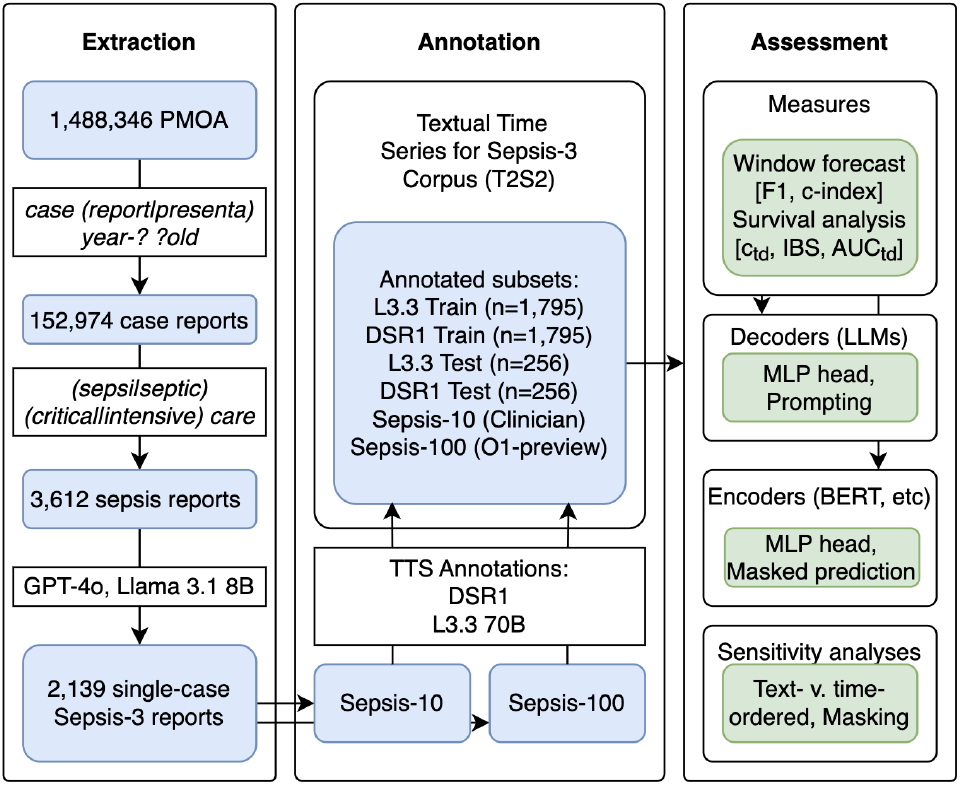
T2S2 forecasting analysis pipeline

### Dataset Extraction

We utilized the PubMed Open Access (PMOA) Subset (as of April 25, 2024) for our analysis. Building on the method described in prior work (Wang and Weiss 2025), the body of each case was extracted by selecting the text between the markers `==== Body` and `==== Ref`, which serve as section delimiters in the PMOA corpus. To maintain relevance, we include only documents containing case-insensitive matches to `(case report|case presenta)` and `year-? ?old`, an approach shown to have higher specificity than PubMed metadata (Noroozizadeh and Weiss 2025).

As demonstrated in prior work of (Noroozizadeh and Weiss 2025), additional filtering was applied to identify potential sepsis-related case reports using the regular expressions `sepsi|septic` and `critical|intensive` care. We then used LLM-based queries to extract key attributes, including sepsis diagnosis, patient count, age, and gender. For this step, both *GPT-4o* and *Llama-3*.*1-8B-Instruct* were employed. Reports describing more than one case were excluded. A case report was retained if either model classified it as involving a Sepsis-3 diagnosis.

This multi-step process yielded a total of 2319 Sepsis-3 case reports. From this set, we randomly selected two subsets for evaluation: a group of 10 reports (sepsis-10), which underwent expert clinical annotation to serve as a gold standard, and a larger group of 100 reports (sepsis-100) as a bronze standard for broader testing based on an alternative annotator (O1-preview; Figure 2).

### Textual Time Series Annotation

Following Noroozizadeh and Weiss (2025), a textual time series case report corpus of Sepsis-3 patients was constructed from the PMOA sepsis reports. A textual time series refers to a sequence of clinical findings, each paired with a timestamp (either absolute or relative to the case presentation time), corresponding to an individual patient. Here, a clinical finding is defined as a free-text expression describing an entity that pertains to or may impact a person’s health. A textual time series differs from conventional approaches to clinical concept annotation (e.g., (Sun, Rumshisky, and Uzuner 2013; Uzuner et al. 2011)) in its extension of the text span to better capture the specificity and contextual meaning of each clinical finding. In our work, the interpretation of a clinical finding differs from the i2b2 concept guidelines in the following respects : (1) Clinical findings are not restricted to single prepositional phrases following a markable entity. For instance, instead of splitting “pain in chest that radiates substernally” into less informative parts, we retain the full phrase as a single, complete finding to preserve its meaning and context. (2) To improve clarity, coordinated phrases are broken into individual findings.

For example, “metastases in the liver and pancreas” becomes “metastasis in the liver” and “metastasis in the pancreas.” We used *DeepSeek-R1-UD-IQ1* and *Llama-3*.*3-70B-Instruct* models to generate clinical textual time series from the 2319 sepsis case reports. We refer to this as **T**extual **T**ime-**S**eries for **S**epsis (T2S2). The exact LLM extraction prompt can be found in Appendix F. Our dataset was split into training (n = 1,795; 80% train, 20% validation), testing T2S2-test (n = 244), and two external validation sets: sepsis-10 (n = 10) and sepsis-100 (n = 90).

### Forecasting Tasks

Using the timelines extracted from the annotations, we defined two primary forecasting tasks and a survival analysis task as follows:

### Event Occurrence Prediction

Given a prefix of the clinical timeline (all events up to a certain time *t*), the model is tasked with predicting whether each of the immediate next *k* events occurs within a specified time horizon. This setup is repeated across multiple time cut-offs to simulate an “online” forecasting scenario, where the model must output a binary label for each of the next *k* events: does this event occur within *h* hours after *t*? Time horizons used include 1 hour, 24 hours (1 day), and 168 hours (1 week). The task is framed as a series of binary classification problems, with one binary decision per event. Evaluation is based on precision, recall, and F1 score, averaged across event positions for each time horizon.

### Temporal Ordering Prediction

This task assesses the model’s ability to reconstruct the correct sequence of future events. At each time cut-off *t*, we extract the next *k* events from the timeline and remove their timestamps. The model must correctly output a permutation over these events that matches their true chronological order. This is framed as a ranking task, evaluated by computing the pairwise concordance between the predicted and true orderings (e.g., proportion of correctly ordered pairs). This tests whether the model can infer temporal progression from unordered event content alone.

### Survival Analysis for Mortality Time

We include a classical survival analysis task to model time until death. Many case reports specify whether the patient died and, if so, when (e.g., “the patient died on hospital day 10”), which enables us to define a time-to-event outcome. For this task, we evaluate models at predefined cut-off times – specifically at 0 hours (admission), 24 hours (1 day), and 168 hours (1 week) – and use the extracted event history up to each cut-off as input. A survival model is trained to predict the probability of survival over time beyond each cut-off. We evaluate using the time-dependent concordance index to measure alignment of the predicted survival times with actual outcomes.

### Modeling Approaches

To evaluate performance on the event forecasting and survival prediction tasks, we implement five modeling paradigms: (i) decoder-only large language models (LLMs) with fine-tuned heads, (ii) prompted LLMs without gradient updates (zero- or few-shot), (iii) encoder-only models with task-specific fine-tuned heads, (iv) encoder-masking models with fine-tuning, and (v) encoder-masking models in zeroshot settings. In Appendix C and D, we provide detailed explanations of implementation details.

#### (i) Fine-tuned LLMs

We apply instruction-tuned decoder-only models from the *LLaMA* and *DeepSeek* families: *Llama-3*.*3-70B-Instruct, Llama-3*.*1-8B-Instruct*, and their distilled variants (Grattafiori et al. 2024; Liu et al. 2024). We also include open-source models with documented training corpora (*OLMO-32B-Instruct* (OLMo et al. 2024), *RedPajama-INCITE-7B-Instruct* (Weber et al. 2024)) to address potential data leakage concerns–notably, RedPajama explicitly excludes PubMed from its training data. Additionally, we evaluate a medically fine-tuned decoder model (*MediPhi-PubMed* (Corbeil et al. 2025)) to assess domain-specific benefits.

Each model is paired with a lightweight multilayer perceptron (MLP) head trained for classification or ranking, depending on the task. The input consists of a text-formatted prefix of events (e.g., a clinical timeline up to time *t*), optionally accompanied by an instruction template. The output layer produces task-specific predictions: binary labels for event occurrence or a permutation over *k* events for ordering. Training is conducted using cross-entropy loss for classification and pairwise ranking loss for ordering.

#### (ii) Prompted LLMs

In the zero- or few-shot setting, we use the same LLM architectures as above, but without any fine-tuning. Instead, we supply structured prompts at inference time to guide the model toward task-specific outputs. Each prompt includes: (1) a system instruction establishing the model’s role (e.g., “You are an expert physician.”), (2) a user instruction describing the prediction task, and (3) one or more input-output examples in a few-shot format. Prompt designs are customized for each task and are detailed in Appendix G. Model outputs are parsed to extract binary labels or ordered lists, and errors in generation or ambiguity are excluded from evaluation.

#### (iii) Fine-tuned Encoder-Only Models

We also evaluate a range of encoder-based models trained end-to-end on each task. Architectures include general-purpose models (*BERT-base-uncased* (Devlin et al. 2019), *RoBERTa-base* (Liu et al. 2019), *DeBERTa-v3-small* (He, Gao, and Chen 2021), *ModernBERT-base/large* (Warner et al. 2024)) and biomedically-pretrained variants (*BioClinical-ModernBERT-base/large* (Sounack et al. 2025)). For each model, we append a task-specific MLP head for event occurrence or ordering prediction. The model input is the same flattened prefix of the event sequence used in other settings, tokenized and formatted according to the respective architecture. These models are trained using standard supervised learning objectives and evaluated on the same metrics as other methods: F1 score for event occurrence, and pairwise concordance for temporal ordering. For survival analysis, the final hidden states are passed into a time-to-event prediction head and evaluated using the time-dependent concordance index (Antolini, Boracchi, and Biganzoli 2005).

#### (iv, v) Encoder-Masking Models

We adapt masked language modeling (MLM) for our temporal reasoning tasks. For event occurrence, models predict masked tokens (“`yes`”/”`no`”) in prompts such as “Will [event] happen within 24 hours? `[MASK]`”. For temporal ordering, models predict “`before`”/”`after`” tokens in prompts comparing event pairs. We evaluate both fine-tuned variants (trained on task-specific objectives) and zero-shot variants (using pretrained MLM capabilities without additional training). This approach leverages the bidirectional context of encoder models while maintaining interpretable predictions through constrained vocabulary.

### Sensitivity Analyses

We conduct sensitivity analyses to examine how the temporal structure and completeness of input sequences affect forecasting performance. These analyses probe the robustness of models to changes in event ordering and to varying levels of missing historical information:

### Time-ordering Strategies

We evaluate two strategies for ordering clinical events within textual time series inputs. In the *text-ordered* setting, events are presented in the order they appear in the original case report narratives, preserving the narrative structure produced during extraction. In the *time-ordered* setting, events are sorted chronologically by their documented occurrence time, enforcing strict temporal alignment. This comparison isolates the effect of narrative sequencing versus explicit temporal ordering on model performance. For text-ordered inputs, to prevent causal leakage, events occurring after the forecast cutoff time *t* are masked for encoder-based models and omitted for decoder-based models.

### Timestep Dropout

To assess robustness to incomplete patient histories, we introduce a *timestep dropout rate* (TDR), defined as the proportion of input timesteps randomly removed prior to inference. This procedure simulates varying degrees of missing clinical documentation that may arise in deployment settings. We vary TDR from 0% (full history) to 90% (severely truncated history) in increments, masking events independently and uniformly at random. This setup enables controlled evaluation of model performance under partial information, probing the extent to which predictive accuracy and event ordering depend on the completeness of historical context.

Together, these analyses characterize model robustness to disruptions in temporal coherence and input completeness – two key challenges in real-world deployment scenarios.

## Results

In this section, we present results for our three main evaluation settings: event forecast within a subsequent time window, temporal ordering of forecasted events, and survival analysis. We then provide sensitivity analyses on the forecasting tasks, examining the effects of temporal ordering and historical context availability.

### Forecasting Tasks

#### Event forecast within next 24 hours: F1 performance

Our results show that encoder-based models outperform decoder-based LLMs in event forecasting across both *DeepSeek-R1* and *Llama-3*.*3-70B* annotations (Table 1). Forecasting performance across all models is shown in Tables A1 and A2 (Appendix A). Encoder models, especially those with a fine-tuned MLP head, achieve substantially higher F1 scores than decoder LLMs, reinforcing the effectiveness of encoder-based representations for forecasting.

**Table 1:** Forecasting performance (event occurrence: F1 and correct event ordering: concordance-index) of the ensuing *k* = 8 events. All models are trained/fine-tuned on time-ordered annotations from either *DeepSeek-R1* or *Llama 3*.*3-70B*. **Bold** values indicate best in category within each column group (refer to Tables A1 and A2 in Appendix A for detailed performance statistics on all models). Performance statistics for all models are presented in A1 and A2, respectively. Abbreviations: F1(1d)-10/100: F1 (1 day) for sepsis-10 and sepsis-100 respectively. c10/100: c-index for sepsis-10 and sepsis-100 respectively.

|  | T2S2 (*DeepSeek-R1*) |  |  |  |  |  |  |  | T2S2 (*Llama 3.3-70B*) |  |  |  |  |  |  |  |
| --- | --- | --- | --- | --- | --- | --- | --- | --- | --- | --- | --- | --- | --- | --- | --- | --- |
|  | F1(1h) | F1(1d) | F1(1w) | c-index | F1(1d)-10 | c10 | F1(1d)-100 | c100 | F1(1h) | F1(1d) | F1(1w) | c-index | F1(1d)-10 | c10 | F1(1d)-100 | c100 |
| **LLM-MLP head** |  |  |  |  |  |  |  |  |  |  |  |  |  |  |  |  |
| DS-L3.3 70B | **0.075** | 0.482 | **0.796** | **0.632** | **0.397** | **0.578** | **0.613** | 0.595 | 0.140 | 0.563 | 0.811 | **0.624** | **0.397** | **0.585** | **0.629** | **0.598** |
| OLMO 32B Instruct | 0.000 | 0.411 | 0.688 | 0.621 | 0.273 | 0.561 | 0.432 | 0.593 | 0.190 | 0.315 | 0.764 | 0.604 | 0.238 | 0.551 | 0.315 | 0.596 |
| RedPajama-INCITE 7B Instruct | 0.000 | 0.352 | 0.651 | 0.618 | 0.282 | 0.572 | 0.471 | 0.594 |  |  |  |  |  |  |  |  |
| MediPhi-PubMed | 0.000 | 0.268 | 0.751 | 0.626 | 0.169 | 0.561 | 0.210 | 0.597 | 0.000 | 0.507 | 0.801 | 0.623 | 0.277 | 0.564 | 0.534 | 0.600 |
| **Prompting** |  |  |  |  |  |  |  |  |  |  |  |  |  |  |  |  |
| L3.3 70B few-shot | **0.095** | **0.380** | 0.652 | – | **0.360** | – | 0.452 | – | 0.064 | **0.460** | 0.729 | – | 0.313 | – | 0.480 | – |
| OLMO 32B few-shot ordinal | 0.013 | 0.315 | 0.651 | – | 0.247 | – | 0.422 | – | 0.046 | 0.413 | 0.647 | – | 0.300 | – | 0.424 | – |
| **Encoder-MLP head** |  |  |  |  |  |  |  |  |  |  |  |  |  |  |  |  |
| ModernBERT-base | 0.306 | **0.576** | 0.877 | 0.646 | 0.428 | 0.562 | 0.607 | 0.558 | 0.257 | 0.645 | **0.903** | 0.594 | 0.431 | 0.590 | 0.626 | 0.565 |
| Bioclinical ModernBERT-base | 0.264 | 0.559 | **0.879** | **0.677** | 0.395 | **0.598** | **0.635** | **0.610** | 0.290 | **0.653** | 0.902 | **0.650** | **0.449** | **0.618** | **0.662** | **0.627** |
| **Encoder-masking-fine-tuned** |  |  |  |  |  |  |  |  |  |  |  |  |  |  |  |  |
| ModernBERT-base | **0.186** | 0.449 | **0.697** | 0.672 | 0.325 | 0.576 | **0.503** | 0.613 | **0.139** | **0.489** | 0.700 | 0.632 | **0.334** | 0.595 | **0.478** | 0.612 |
| Bioclinical ModernBERT-base | 0.166 | 0.450 | 0.692 | **0.676** | 0.336 | 0.604 | 0.499 | **0.655** | 0.129 | 0.481 | 0.733 | 0.653 | 0.331 | **0.615** | 0.475 | **0.658** |
| **Encoder-masking-zeroshot** |  |  |  |  |  |  |  |  |  |  |  |  |  |  |  |  |
| ModernBERT-base | 0.169 | **0.468** | **0.804** | 0.496 | **0.358** | 0.501 | **0.511** | 0.499 | 0.132 | **0.525** | **0.840** | 0.556 | **0.352** | 0.503 | 0.511 | 0.501 |
| Bioclinical ModernBERT-base | **0.246** | 0.458 | 0.614 | **0.498** | 0.301 | 0.509 | 0.457 | 0.498 | **0.197** | 0.460 | 0.533 | 0.499 | 0.299 | 0.520 | 0.465 | 0.499 |

Among decoder models, we observe varied performance patterns. *RedPajama-INCITE-7B-Instruct*, with documented training that excludes PubMed, achieves 0.352 F1 at 24h— better than some decoder models like *MediPhi-PubMed* (0.268) and within the range of decoder performance, though below top performers like *DeepSeek-Llama-3*.*3-70B* (0.482). *OLMO-32B-Instruct* performs competitively (0.411), suggesting that open models without biomedical pretraining can still achieve reasonable results for temporal forecasting tasks.

Among encoder models, both general-purpose and biomedically-pretrained variants show strong performance. While *ModernBERT-base* achieves slightly higher F1 scores on internal test sets (0.576 vs 0.559), *BioClinical-ModernBERT-base* demonstrates superior concordance (0.677 vs 0.646 for DeepSeek-R1) and notably better generalization to external datasets, with F1 scores of 0.635 and 0.662 on sepsis-100 compared to 0.607 and 0.626 for the general-purpose variant. This suggests that while both ModernBERT architectures are effective, biomedical pretraining particularly enhances temporal reasoning and cross-dataset generalization.

Different training strategies yield distinct performance patterns. The MLP head fine-tuning approach consistently outperforms both fine-tuned masking and zero-shot masking models, particularly in long-horizon forecasting. While *ModernBERT-base* achieves strong F1 scores, *BioClinical-ModernBERT-base* demonstrates the best overall performance when considering both accuracy and generalization. Zero-shot masking models, particularly standard *BERT* and *RoBERTa*, fail to make meaningful predictions at the 1-hour mark. However, *ModernBERT* variants exhibit relatively better zero-shot performance, with *BioClinical-ModernBERT-base* achieving notably higher scores in zeroshot settings (0.246 F1 at 1h), indicating that biomedical pretraining improves out-of-the-box temporal reasoning.

Performance trends remain consistent across forecasting windows. F1 scores improve as the window increases, with 1-hour predictions being most challenging and 168-hour predictions yielding highest scores. Models perform worse on external validation sets than internal T2S2 test sets, though *BioClinical-ModernBERT* models show the smallest performance degradation, maintaining their advantage in cross-dataset generalization.

#### Temporal ordering of forecasted events: concordance

Encoder models consistently outperform decoders in correctly ranking the order of upcoming events, as measured by concordance (c-index; Table 1; complete results in Tables A1 and A2 for *DeepSeek-R1* and *Llama-3*.*3-70B* annotations, respectively, with full details in Appendix A. Decoder models with potential biomedical exposure achieve modest concordance (0.618-0.632), while *RedPajama-INCITE-7B-Instruct* without PubMed data in its training corpus performs similarly, suggesting domain-specific knowledge is less critical here than for event prediction. The comparable performance of *RedPajama-INCITE-7B-Instruct* to other general models (with access to PubMed in their pretraining) also provides evidence that the T2S2 prediction task is distinct from raw PubMed text and assuages causal leakage concerns.

Biomedical pretraining in encoder models also demonstrates clear advantages, with *BioClinical-ModernBERT-base* attaining the highest concordance (0.677 with MLP head, 0.676 fine-tuned masking) and robust performance across datasets. MLP-head training yields superior F1 scores, while fine-tuned masking excels in concordance. *BioClinical-ModernBERT-base* maintains strong generalization on external datasets (c-index *>*0.60), suggesting that biomedical pretraining provides stability in temporal reasoning across dataset shifts.

#### Survival analysis: model performance across timepoints and cohorts

Table 2 presents time-dependent concordance across evaluation cohorts (*T2S2, sepsis-10, sepsis-100*) at multiple observation windows (0h, 24h, 168h), selecting models based on validation concordance. General models, such as *RedPajama-INCITE-7B-Instruct*, show strong survival prediction capabilities even without biomedical exposure (0.76 concordance at 168h, sepsis-100). Mean-while, biomedical encoders like *BioClinical-ModernBERT-base* consistently perform well, particularly early (0h).

**Table 2:**
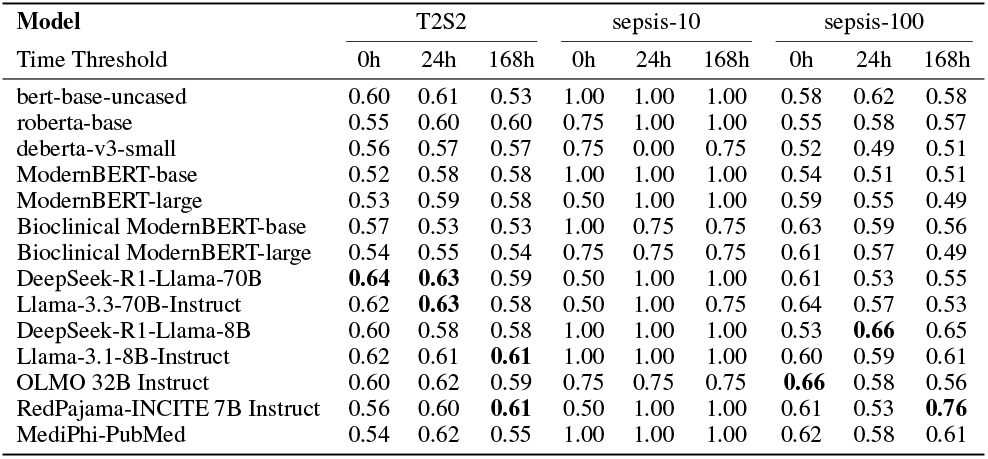
Survival analyses evaluated using time-dependent concordance index for each model across *DeepSeek-R1* test set annotations (*T2S2, Sepsis-10, Sepsis-100*), evaluated at three observation time thresholds (0h, 24h, and 168h).

| Model | T2S2 |  |  | sepsis-10 |  |  | sepsis-100 |  |  |
| --- | --- | --- | --- | --- | --- | --- | --- | --- | --- |
|  | 0h | 24h | 168h | 0h | 24h | 168h | 0h | 24h | 168h |
| bert-base-uncased | 0.60 | 0.61 | 0.53 | 1.00 | 1.00 | 1.00 | 0.58 | 0.62 | 0.58 |
| roberta-base | 0.55 | 0.60 | 0.60 | 0.75 | 1.00 | 1.00 | 0.55 | 0.58 | 0.57 |
| deberta-v3-small | 0.56 | 0.57 | 0.57 | 0.75 | 0.00 | 0.75 | 0.52 | 0.49 | 0.51 |
| ModernBERT-base | 0.52 | 0.58 | 0.58 | 1.00 | 1.00 | 1.00 | 0.54 | 0.51 | 0.51 |
| ModernBERT-large | 0.53 | 0.59 | 0.58 | 0.50 | 1.00 | 1.00 | 0.59 | 0.55 | 0.49 |
| Bioclinical ModernBERT-base | 0.57 | 0.53 | 0.53 | 1.00 | 0.75 | 0.75 | 0.63 | 0.59 | 0.56 |
| Bioclinical ModernBERT-large | 0.54 | 0.55 | 0.54 | 0.75 | 0.75 | 0.75 | 0.61 | 0.57 | 0.49 |
| DeepSeek-R1-Llama-70B | **0.64** | **0.63** | 0.59 | 0.50 | 1.00 | 1.00 | 0.61 | 0.53 | 0.55 |
| Llama-3.3-70B-Instruct | 0.62 | **0.63** | 0.58 | 0.50 | 1.00 | 0.75 | 0.64 | 0.57 | 0.53 |
| DeepSeek-R1-Llama-8B | 0.60 | 0.58 | 0.58 | 1.00 | 1.00 | 1.00 | 0.53 | **0.66** | 0.65 |
| Llama-3.1-8B-Instruct | 0.62 | 0.61 | **0.61** | 1.00 | 1.00 | 1.00 | 0.60 | 0.59 | 0.61 |
| OLMO 32B Instruct | 0.60 | 0.62 | 0.59 | 0.75 | 0.75 | 0.75 | **0.66** | 0.58 | 0.56 |
| RedPajama-INCITE 7B Instruct | 0.56 | 0.60 | **0.61** | 0.50 | 1.00 | 1.00 | 0.61 | 0.53 | **0.76** |
| MediPhi-PubMed | 0.54 | 0.62 | 0.55 | 1.00 | 1.00 | 1.00 | 0.62 | 0.58 | 0.61 |

Predictive performance does not universally improve with more observation time. *T2S2* cohort models often peak at earlier windows, while *sepsis-10* experiences ceiling effects. Instruction-tuned decoders excel overall, yet biomedical encoders remain competitive, particularly at admission (0h). Survival prediction thus benefits from both larger decoder models and domain-specific pretraining, each excelling in different temporal contexts.

### Sensitivity analysis

#### Time-Ordered versus Text-Ordered Training

Table 3 shows time-ordering generally improves concordance, particularly for *DeepSeek-R1* annotations, while F1 scores show mixed results. Text-ordering sometimes yields better external dataset performance with *Llama-3*.*3-70B*, suggesting that preserving the narrative sequence can support accurate classification even when it does not follow temporal order.

**Table 3:** Comparison of text-ordered and time-ordered training for *DeepSeek-R1* and *Llama 3*.*3-70B* annotations on F1 scores and c-index values.

|  | F1(1h) | F1(1d) | F1(1w) | c-index | F1(1d)-10 | c10 | F1(1d)-100 | c100 |
| --- | --- | --- | --- | --- | --- | --- | --- | --- |
| **T2S2 (*DeepSeek-R1*)** |  |  |  |  |  |  |  |  |
| Text-ordered | 0.290 | 0.569 | 0.876 | 0.659 | 0.361 | 0.598 | 0.615 | 0.609 |
| Time-ordered | 0.306 | 0.576 | 0.877 | 0.672 | 0.428 | 0.576 | 0.607 | 0.613 |
| **T2S2 (*Llama 3.3-70B*)** |  |  |  |  |  |  |  |  |
| Text-ordered | 0.311 | 0.648 | 0.898 | 0.586 | 0.423 | 0.526 | 0.662 | 0.541 |
| Time-ordered | 0.257 | 0.645 | 0.903 | 0.632 | 0.382 | 0.595 | 0.616 | 0.612 |

These results highlight a trade-off between temporal alignment and narrative coherence for different tasks.

#### Impact of timestep dropout on F1 and concordance

Tables 4 and A3 (Appendix B) report the effect of timestep dropout on classification and ranking performance. F1 scores decrease monotonically with increasing TDR, with relatively mild degradation up to 30–60% dropout and sharper declines thereafter. For example, in the *DeepSeek-R1* annotations, T2S2/sepsis-10/sepsis-100 drop from 0.576/0.428/0.607 at TDR = 0% to 0.423/0.200/0.510 at TDR = 90%, with sepsis-10 showing the largest relative decline. In contrast, concordance (c-index) remains stable across dropout levels (0.661–0.678), indicating that ranking-based evaluations are less sensitive to history truncation. These trends are consistent for *Llama-3*.*3* annotations (Appendix B), suggesting that ranking objectives capture temporal relationships that are more robust to missing historical information than binary classification accuracy of predicting whether an events happens within a time window.

**Table 4:**
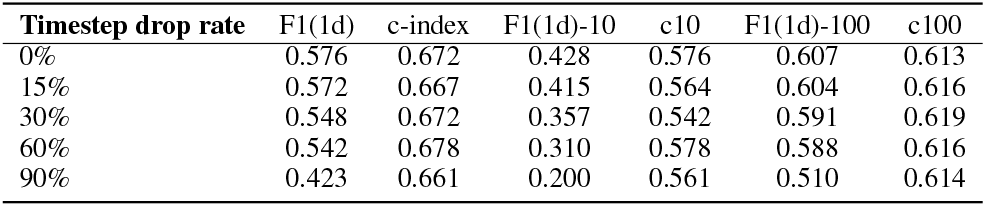
Effect of Randomly Masking History (*DeepSeek-R1* annotations) on F1 and Concordance.

## Discussion

Our findings highlight the superiority of encoder-based models over decoder-based LLMs for event forecasting, emphasizing the limitations of autoregressive models in structured prediction tasks. Among encoder models, *BioClinical-ModernBERT-base* consistently performs best, achieving superior concordance (0.677 vs 0.646) and better external generalization despite slightly lower internal F1 scores than general-purpose *ModernBERT-base*. MLP-head fine-tuning excels in F1 scores while fine-tuned masking models achieve higher concordance, particularly in long-term forecasting. This suggests that classification and ranking tasks benefit from distinct optimization strategies.

The consistent advantage of biomedically-pretrained models across tasks deserves special attention. While general-purpose models achieve strong internal performance, biomedical pretraining provides crucial benefits for real-world deployment: *BioClinical-ModernBERT* variants show the smallest performance degradation on external datasets and maintain higher zero-shot capabilities (0.246 F1 at 1h vs near-zero for standard BERT/RoBERTa). This suggests that domain-specific pretraining enhances not just accuracy but also robustness and generalization—critical factors when considering clinical deployment where distribution shifts are common.

The gradual improvement of F1 scores over longer time horizons indicates that event patterns become more predictable over time, whereas short-term forecasting remains challenging due to higher variability. Performance drops on external validation datasets highlight generalization challenges, though *ModernBERT* models exhibit relative robustness. For survival analysis, instruction-tuned LLMs like *Llama-3*.*3-70B-Instruct* outperform traditional transformer baselines, with several models achieving peak concordance at early timepoints rather than with extended observation.

Our sensitivity analysis reveals several trade-offs: timeordered training generally improves concordance while text-ordering can yield better F1 scores on externally-annotated datasets. Robustness experiments show F1 scores degrade significantly beyond 60% timestep dropout, while concordance remains stable, indicating event ranking is less sensitive to partial history than event classification.

### Methodological Contributions and Potential Impact

Our framework demonstrates how narrative clinical texts can be systematically converted into structured temporal representations for forecasting tasks. The ability to extract temporally structured insights from unstructured clinical text could potentially support clinical decision-making, particularly in settings with limited access to structured data or specialized expertise.

Importantly, while consumers frequently consult LLMs about health risk via prompting in chats, this study demonstrates that the prompting approach performs substantially worse in risk prediction than alternative approaches, at least with respect to precision/recall/F1, with prompted LLMs achieving at best 0.460 F1 at 24h compared to 0.653 for fine-tuned encoders. Our study highlights several alternative approaches and characterizes their relative performance strengths and weaknesses. In addition, our finding that concordance remains stable with 60% missing context suggests robustness to incomplete documentation scenarios, which highlights the degradation pattern in performance as it relates to context availability.

Our approach also demonstrates that our language model systems can reliably capture the temporal reasoning that clinicians use in practice based on their clinical documentation. However, additional work would be required to validate performance on real-time clinical data and demonstrate measurable impact on patient outcomes in deployment. Further discussion on societal impact of our work is in Appendix I.

### Limitations and Future Directions

Our study has important limitations to consider. First, the pipeline relies on case reports from the PubMed Open Access (PMOA) corpus, which often reflect rare or atypical presentations and differ from routine clinical notes (e.g., progress notes, discharge summaries), potentially limiting the model’s generalizability to real-world health system texts. Compared to routine clinical notes, case reports offer rich, temporally structured narratives with explicit clinical rationale, making them especially useful for evaluating model performance under sparse documentation. This study develops a general framework for textual time series forecasting, using case reports as an interpretable, temporally rich foundation for method development with potential extensions to real-world clinical corpora like MIMIC discharge summaries.

Second, while we focused on sepsis due to its clinical relevance and prevalence in case reports, our framework is fundamentally disease-agnostic. Preliminary results from over 125K PMOA case reports across various diagnostic conditions show promising generalization, with future work planned on broader diagnostic categories and real-world corpora such as MIMIC-IV discharge summaries.

Third, despite efforts to mitigate causal leakage during training and evaluation (e.g., timestamp extraction, temporal masking, and use of external test sets), pretrained language models may have been exposed to PMOA content. Although these models were not trained on our derived (event, time) sequences or temporally reordered data, it is possible that the original case report narratives were included in their pretraining data. Our pipeline applies a series of transformations—including temporal reordering and masking of future events beyond a prediction cutoff—which fundamentally alter the input representation and enforce a causal structure that is not present in the original documents. This creates a distinct prediction setup that requires models to reason over partial, temporally grounded inputs rather than complete narratives. To further assess the impact of pretraining exposure, we include results from *RedPajama-INCITE-7B-Instruct*, an open-source model with a fully documented, PubMed-free training corpus. Its comparable performance to other decoder models (0.352 F1 at 24h) and similar concordance (0.618) to models potentially exposed to PubMed provides evidence that our T2S2 prediction task is sufficiently distinct from raw PubMed text, helping to mitigate causal leakage concerns. A detailed discussion on the scope of using open-source models for avoiding potential data leakage has been presented in Appendix H.

Finally, our analysis is limited by the modest size of the high-cost manual annotations collected as a gold standard to calibrate and validate our LLM extraction pipeline. Expanding gold-standard validation across diverse diagnoses could assess the generalizability of our approach and aid in evaluating model robustness, and is a direction for future work.

## Data Availability

We utilized case reports from the publicly available PubMed Open Access (PMOA) Subset (as of April 25, 2024) for our analysis.

## Acknowledgments

This research was supported in part by the Division of Intramural Research (DIR) of the National Library of Medicine (NLM), National Institutes of Health. This work utilized the computational resources of the NIH HPC Biowulf cluster. S. N. was supported by Carnegie Mellon University TCS Presidential Fellowship, and Natural Sciences and Engineering Research Council of Canada (NSERC) PGS-D award. S.N was also supported in part by an appointment to the National Library of Medicine Research Participation Program administered by the Oak Ridge Institute for Science and Education (ORISE) through an interagency agreement between the U.S. Department of Energy (DOE) and the National Library of Medicine, National Institutes of Health. ORISE is managed by ORAU under DOE contract number DE-SC0014664. All opinions expressed in this paper are the authors’ and do not necessarily reflect the policies and views of NIH, NLM, DOE, or ORAU/ORISE.

## Supplementary Material

This supplementary material provides detailed information organized as follows:

- **Appendix A**: Complete forecasting performance tables for all models on DeepSeek-R1 and Llama-3.3-70B annotations
- **Appendix B**: Sensitivity analysis of masking history on Llama-3.3-70B annotations
- **Appendix C**: Technical details of encoder-based forecasting methods, including MLP-head and masking approaches
- **Appendix D**: Survival modeling framework using language model embeddings and the hyperparameters used
- **Appendix E**: Computing infrastructure used for experiments, including hardware specifications, memory requirements, operating system, and software environment
- **Appendix F**: LLM prompt template for extracting clinical time series from case reports
- **Appendix G**: Prompting strategies for zero-shot and few-shot forecasting
- **Appendix H**: Analysis of open-source models to address data leakage concerns
- **Appendix I**: Broader societal impact and ethical considerations

### A Forecasting performance on *DeepSeek-R1* and *Llama-3*.*3-70B* textual time-series annotations

Tables A1 and A2 present the forecasting performance of our models on textual time-series annotations generated by *DeepSeek-R1* and *Llama-3*.*3-70B*, respectively. We report standard forecasting metrics across multiple evaluation settings to provide a comprehensive view of model performance.

### B Masking history: *Llama-3*.*3-70B* textual time-series annotations

Table A3 examines the impact of randomly masking portions of the input history on model performance when using *Llama-3*.*3-70B*-generated annotations. We evaluate performance across three datasets (T2S2-L33, Sepsis-10, and Sepsis-100) using F1 scores for prediction of next *k* events happening with the next day and concordance index (cindex) correctly ordering the next *k* events. As the proportion of masked history increases, we observe a general decline in F1 performance, particularly at higher masking rates (e.g., 90%), suggesting that access to historical context is important for accurate forecasting. Interestingly, the concordance index remains relatively stable, indicating the model’s robustness in ranking the next *k* events with partial input histories.

### C Textual Time Series Forecasting Framework of Encoder Models

We develop a general framework for predictive modeling over textual time series of the encoder-only models, designed to accommodate the irregular, event-based structure common to clinical case reports. As highlighted Section, each data instance consists of a sequence of clinical events, where each event is a free-text string describing a clinical finding, intervention, or diagnosis, paired with a timestamp in hours relative to the time of admission. The format resembles a two-column table, where the first column contains textual clinical events and the second column contains integer-valued timestamps.

#### Input Preprocessing and Timeseries Construction

From each case report, we construct multiple training examples for sequence forecasting. For a fixed forecast horizon parameter *K*, we generate overlapping windows of historical events and forecast targets. Specifically, for each case report with *L* rows, we construct a sequence of up to *L − K* forecasting examples. Each example is structured as follows:

- A **history segment** 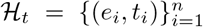, consisting of clinical events *e*_*i*_ occurring at timestamps *t*_*i*_ *≤ t*.
- A **forecast target** 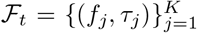, consisting of the next *K* events after *t*.

To preserve the temporal coherence of events, we treat all events with the same timestamp as an atomic unit. That is, if multiple events share the same timestamp, they are included together in the history or forecast block, and the next forecasting window is only advanced after the full timestamp block has been consumed.

#### Model Input Representation

Each training example is serialized into a transformer-compatible input by linearizing the historical events and forecast events. Importantly, we preserve both temporal and textual content by encoding each event in the form “`t:e`” where `t` is the timestamp and `e` is the event string. The input to the language model takes the form:

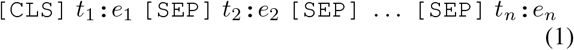

This is followed by additional target-specific components, as described below.

#### Language Models

To test the generality of our framework, we experiment with a variety of pre-trained encoderonly language models, including:

- *bert-base-uncased* (BERT; Devlin et al., 2018)
- *roberta-base* (Liu et al., 2019)
- *microsoft/deberta-v3-small* (He et al., 2021)
- *answerdotai/ModernBERT-base* and
- *answerdotai/ModernBERT-large*
- *thomas-sounack/BioClinical-ModernBERT-base* and
- *thomas-sounack/BioClinical-ModernBERT-large*

**Table A1:** Forecasting performance (event occurrence: F1 and correct event ordering: concordance-index) of the ensuing *k* = 8 events. Models *italicized* represent the open-source and medically fine-tuned models. All models are trained/fine-tuned on time-ordered annotations from *DeepSeek-R1*. **Bold** indicates best in that particular category amongst all models.

|  | T2S2 (DeepSeek R1) |  |  |  | sepsis-10 |  | sepsis-100 |  |
| --- | --- | --- | --- | --- | --- | --- | --- | --- |
|  | F1(1h) | F1(1d) | F1(1w) | c-index | F1(1d) | c-index | F1(1d) | c-index |
| **LLM-MLP head** |  |  |  |  |  |  |  |  |
| DS-L3.3 70B | **0.075** | 0.482 | **0.796** | **0.632** | **0.397** | **0.578** | **0.613** | 0.595 |
| L3.3 70B | 0.000 | 0.295 | 0.764 | 0.631 | 0.312 | 0.569 | 0.404 | 0.597 |
| DS-L3.1 8B | 0.000 | **0.492** | 0.650 | 0.630 | 0.381 | 0.566 | 0.601 | 0.600 |
| L3.1 8B | 0.000 | 0.355 | 0.650 | 0.630 | 0.114 | 0.575 | 0.442 | **0.604** |
| *OLMO 32B Instruct* | 0.000 | 0.411 | 0.688 | 0.621 | 0.273 | 0.561 | 0.432 | 0.593 |
| *RedPajama-INCITE 7B Instruct* | 0.000 | 0.352 | 0.651 | 0.618 | 0.282 | 0.572 | 0.471 | 0.594 |
| *MediPhi-PubMed* | 0.000 | 0.268 | 0.751 | 0.626 | 0.169 | 0.561 | 0.210 | 0.597 |
| **Prompting** |  |  |  |  |  |  |  |  |
| L3.3 70B 0-shot | 0.085 | 0.368 | 0.560 | – | 0.236 | – | 0.452 | – |
| L3.3 70B 0-shot ordinal | 0.082 | 0.352 | 0.646 | – | 0.289 | – | **0.483** | – |
| L3.3 70B few-shot | **0.095** | **0.380** | 0.652 | – | **0.360** | – | 0.452 | – |
| L3.3 70B few-shot ordinal | 0.063 | 0.356 | 0.682 | – | 0.313 | – | 0.480 | – |
| *OLMO 32B 0-shot* | 0.018 | 0.333 | 0.652 | – | 0.322 | – | 0.430 | – |
| *OLMO 32B 0-shot ordinal* | 0.006 | 0.323 | **0.685** | – | 0.325 | – | 0.443 | – |
| *OLMO 32B few-shot* | 0.014 | 0.316 | 0.630 | – | 0.219 | – | 0.423 | – |
| *OLMO 32B few-shot ordinal* | 0.013 | 0.315 | 0.651 | – | 0.247 | – | 0.422 | – |
| **Encoder-MLP head** |  |  |  |  |  |  |  |  |
| BERT | 0.264 | 0.550 | 0.871 | 0.501 | **0.441** | 0.498 | 0.614 | 0.501 |
| RoBERTa | 0.250 | 0.562 | 0.866 | 0.500 | 0.405 | 0.500 | 0.614 | 0.500 |
| DeBERTa-small | 0.262 | 0.570 | 0.871 | 0.601 | 0.382 | 0.498 | 0.617 | 0.556 |
| ModernBERT-base | 0.306 | **0.576** | 0.877 | 0.646 | 0.428 | 0.562 | 0.607 | 0.558 |
| ModernBERT-large | 0.277 | 0.568 | 0.874 | 0.598 | 0.391 | 0.539 | 0.602 | 0.553 |
| *Bioclinical ModernBERT-base* | 0.264 | 0.559 | **0.879** | **0.677** | 0.395 | **0.598** | **0.635** | **0.610** |
| *Bioclinical ModernBERT-large* | **0.313** | 0.562 | 0.868 | 0.671 | 0.399 | 0.491 | 0.618 | 0.607 |
| **Encoder-masking-fine-tuned** |  |  |  |  |  |  |  |  |
| BERT | 0.166 | 0.432 | 0.690 | 0.648 | 0.323 | 0.554 | 0.482 | 0.600 |
| RoBERTa | 0.169 | 0.416 | 0.688 | 0.646 | 0.313 | 0.539 | 0.457 | 0.609 |
| DeBERTa-small | 0.175 | 0.435 | 0.695 | 0.671 | 0.325 | 0.579 | 0.492 | 0.617 |
| ModernBERT-base | **0.186** | 0.449 | **0.697** | 0.672 | 0.325 | 0.576 | **0.503** | 0.613 |
| ModernBERT-large | 0.177 | **0.460** | 0.683 | 0.663 | **0.342** | 0.615 | 0.496 | 0.632 |
| *Bioclinical ModernBERT-base* | 0.166 | 0.450 | 0.692 | **0.676** | 0.336 | 0.604 | 0.499 | **0.655** |
| *Bioclinical ModernBERT-large* | 0.165 | 0.422 | 0.696 | 0.652 | 0.325 | **0.626** | 0.425 | 0.603 |
| **Encoder-masking-zeroshot** |  |  |  |  |  |  |  |  |
| BERT | 0.000 | 0.000 | 0.000 | 0.495 | 0.000 | 0.513 | 0.000 | 0.494 |
| RoBERTa | 0.049 | 0.207 | 0.375 | 0.500 | 0.137 | 0.500 | 0.318 | 0.500 |
| DeBERTa-small | 0.170 | 0.460 | 0.323 | 0.497 | 0.268 | **0.512** | 0.376 | **0.509** |
| ModernBERT-base | 0.169 | **0.468** | **0.804** | 0.496 | 0.358 | 0.501 | **0.511** | 0.499 |
| ModernBERT-large | 0.133 | 0.349 | 0.534 | 0.494 | 0.271 | 0.509 | 0.277 | 0.502 |
| *Bioclinical ModernBERT-base* | **0.246** | 0.458 | 0.614 | **0.498** | 0.301 | 0.509 | 0.457 | 0.498 |
| *Bioclinical ModernBERT-large* | 0.244 | 0.446 | 0.596 | 0.479 | **0.364** | 0.502 | 0.416 | 0.497 |

**Table A2:**
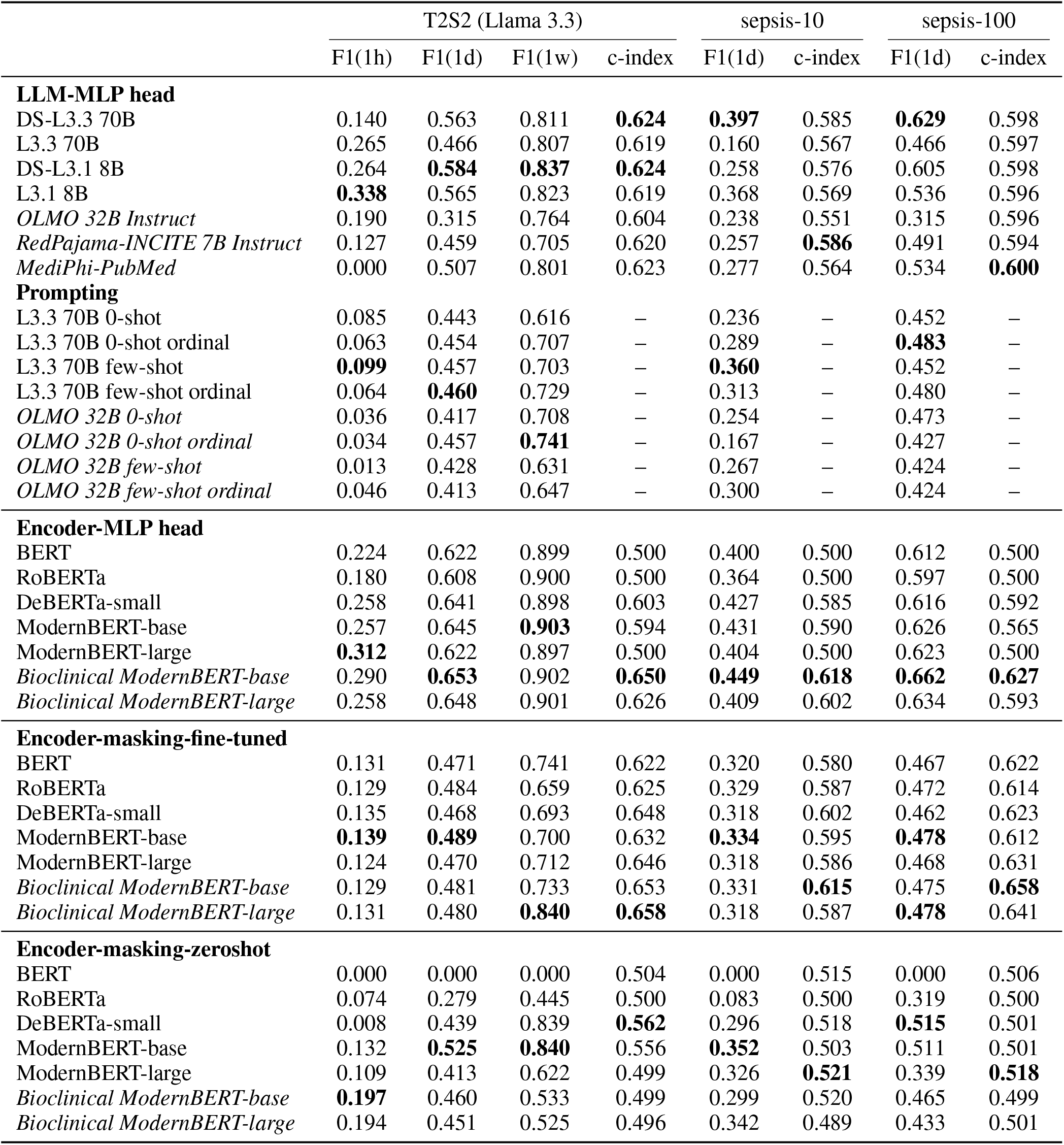
Forecasting performance (event occurrence: F1 and correct event ordering: concordance-index) of the ensuing *k* = 8 events. Models *italicized* represent the open-source and medicallty fine-tuned models. **All models are trained/fine-tuned on time-ordered annotations from *Llama-3*.*3-70B***.

**Table A3:** Effect of Randomly Masking History (*Llama-3*.*3-70B* annotations) on F1 and Concordance. **Abbreviations**: F1(1d)-10/100: F1 (1 day) for sepsis-10 and sepsis-100 respectively. c10/100: c-index for sepsis-10 and sepsis-100 respectively.

| Timestep drop rate | F1(1d) | c-index | F1(1d)-10 | c10 | F1(1d)-100 | c100 |
| --- | --- | --- | --- | --- | --- | --- |
| 0% | 0.645 | 0.632 | 0.431 | 0.595 | 0.626 | 0.612 |
| 15% | 0.628 | 0.635 | 0.422 | 0.592 | 0.624 | 0.610 |
| 30% | 0.634 | 0.634 | 0.423 | 0.586 | 0.621 | 0.598 |
| 60% | 0.604 | 0.634 | 0.391 | 0.577 | 0.605 | 0.613 |
| 90% | 0.531 | 0.622 | 0.328 | 0.546 | 0.534 | 0.607 |

Each model is used with its corresponding tokenizer. For models with SentencePiece tokenization (e.g., DeBERTa), we use the appropriate fast tokenizer such as *DebertaV2Tokenizer* to ensure compatibility. All inputs are truncated from the left (i.e., keeping the most recent tokens) if they exceed the model’s context window (typically 512 tokens).

#### Prediction Tasks

We define two primary prediction tasks over ℱ_*t*_, each formulated as a binary classification problem.

1. **Event Ordering (Concordance) Task**. The goal in this task is to predict the correct temporal ordering between future events. For each example, we generate all *comparable pairs* of events in ℱ_*t*_, defined as pairs (*f*_*i*_, *f*_*j*_) where *τ*_*i*_ ≠ *τ*_*j*_. For each pair, we form a binary classification example as follows:
  - **Input:** The serialized history segment followed by:

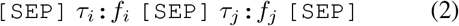
  - **Label:** *y* = 1 if *τ*_*i*_ *< τ*_*j*_, else *y* = 0 The model output is taken from the `[CLS]` token, passed through a linear classification head, and softmaxed to produce a binary probability. We optimize cross-entropy loss across all pairwise comparisons. At evaluation time, we report the concordance index (c-index), which measures the proportion of correctly predicted pairs among all comparable pairs.
2. **Time-Window Classification Task**. This task evaluates whether each forecast event occurs within a fixed prediction window *H* hours of the last historical event. For each *f*_*j*_ *∈* ℱ_*t*_, we generate an input:
  - **Input:** The serialized history segment followed by:

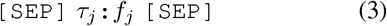
  - **Label:** *y* = 1 if *τ*_*j*_ *− t*_*n*_ *≤ H*, where *t*_*n*_ is the timestamp of the last historical event. This is again treated as a binary classification task over the `[CLS]` embedding, using a linear layer and softmax. We report macro-averaged F1 scores over all prediction examples.

#### Masked Language Modeling Approaches

In addition to the MLP-head approach described above, we implement masked language modeling (MLM) variants that leverage the pretrained masked token prediction capabilities of encoder models. These approaches reformulate our forecasting tasks as cloze-style problems:

1. **Masked Event Occurrence Prediction**. For the time-window classification task, we construct prompts that include a `[MASK]` token whose prediction indicates whether an event occurs within the specified horizon:
  - **Input:** History formatted as: `[CLS]` *h*_1_ `[SEP]` *h*_2_ `[SEP]` … `[SEP]` Will “*e*_*j*_” happen within *H* hours? `[MASK] [SEP]`
  - **Target:** The model predicts either “`yes`” or “`no`” at the `[MASK]` position We constrain the output vocabulary to only these two tokens and compute cross-entropy loss over their logits. For fine-tuned variants, we train the entire model to optimize this masked prediction task. For zero-shot variants, we directly use the pretrained model’s predictions without additional training.
2. **Masked Temporal Ordering**. For the concordance task, we predict relational tokens that indicate temporal ordering:
  - **Input:** History followed by: `[SEP]` *e*_1_ `[MASK]` *e*_2_ `[SEP]`
  - **Target:** The model predicts “`before`” if *e*_1_ occurs before *e*_2_, else “`after`”

Again, we restrict predictions to these two tokens. This formulation allows the model to leverage its pretrained understanding of temporal relationships encoded during MLM pretraining.

Note that in our dataset, we randomize the relative ordering of *e*_1_ and *e*_2_ each time to avoid introducing positional biases that could lead the model to rely on superficial cues rather than genuine temporal reasoning.

#### Implementation Details

For both tasks, we extract the logits at the `[MASK]` position and apply a linear projection to the target vocabulary size (2 tokens). The key advantage of this approach is that it can leverage pretrained masked language modeling capabilities, potentially requiring less task-specific training. We evaluate both fine-tuned versions (which update all model parameters) and zero-shot versions (which use frozen pretrained weights).

#### Training and Evaluation

Each task is trained separately using AdamW with linear learning rate decay and early stopping based on validation loss. We save the model checkpoint with the best validation performance. During inference, we reload this checkpoint and evaluate on a held-out test set (which is obtained from textual time series of unseen case reports).

For both tasks, we extract softmax probabilities rather than hard labels to allow for threshold calibration and ROC-based analyses. All training is conducted using PyTorch and HuggingFace Transformers.

### D Survival Modeling with Language Model

#### Embeddings

To evaluate the prognostic value of textual information encoded in large language models (LLMs), we adopt a two-stage framework: (1) extraction of fixed-dimensional sequence embeddings from various pre-trained LLMs, and (2) downstream survival modeling using these embeddings as covariates.

In the first stage, we process each textual time series (which is converted to a textual context of *“(time) clinical event [SEP]* … *[SEP] (time) clinical event [SEP]”*) using a suite of LLMs to obtain dense vector representations that summarize the content of the input sequence. For models belonging to the encoder family, including *bert-base-uncased, roberta-base, deberta-v3-small, ModernBERT-base*, and *ModernBERT-large*, we extract the final hidden state corresponding to the `[CLS]` token, which is conventionally used to represent the entire sequence in classification tasks. This token-specific embedding serves as a compact, sequence-level representation.

For decoder-based models that do not utilize a dedicated `[CLS]` token, including *DeepSeek-R1-Distill-Llama-70B, Llama-3*.*3-70B-Instruct, DeepSeek-R1-Distill-Llama-8B*, and *Llama-3*.*1-8B-Instruct*, we compute the mean-pooled embedding over the last hidden states of all non-padding tokens. This pooling strategy yields a fixed-length vector that captures the overall semantic content of the input while mitigating the impact of padding artifacts.

In the second stage, these embedding vectors are used as input covariates to three survival models, each designed to capture time-to-event dynamics in different ways:

- **Random Survival Forest (RSF):** A nonparametric ensemble method based on decision trees, capable of handling complex interactions and high-dimensional inputs. The model is tuned over the number of trees (`n_estimators`), the minimum number of samples required to split an internal node (`min_samples_split`), and the minimum number of samples at a leaf node (`min_samples_leaf`); see Table A4 for search ranges.
- **DeepSurv:** A neural-network generalization of the Cox proportional hazards model. The architecture comprises fully connected layers with varying widths (`num_nodes`) and dropout rates; see Table A4 for search ranges. The number of training epochs is fixed.
- **DeepHit:** A multi-task neural network that jointly models the discrete hazard and survival functions. As with DeepSurv, we perform a grid search over hidden layer sizes and dropout rates while keeping the number of epochs fixed; see Table A4.

**Table A4:** Hyperparameter search ranges for survival models.

| Model | Hyperparameters |
| --- | --- |
| RSF | `n_estimators` $\in \{20, 50, 100, 200, 500, 1000\}$<br>`min_samples_split` $\in \{2, 10, 20, 50, 100\}$<br>`min_samples_leaf` $\in \{1, 5, 10, 20\}$ |
| DeepSurv/DeepHit | `num_nodes` $\in \{32, 64, 128, 256, 512, 768, 1024, 1200, 1536, 1800, 2048, 3072, 4096\}$<br>`dropout` $\in \{0.1, 0.2, 0.4, 0.5\}$ |

Hyperparameter tuning for each survival model is performed using a validation split from the training set. For each combination of language model and survival model, we conduct grid search over the respective hyperparameter space, selecting the configuration that maximizes the time-dependent concordance index (Antolini, Boracchi, and Biganzoli 2005) on the validation set. Final evaluation is conducted on held-out test sets (*t2s2_test, sepsis-10, sepsis-100*), and results are aggregated across survival metrics to assess model performance.

This framework allows for a systematic evaluation of how different pre-trained LLMs, and their associated embedding strategies, contribute to downstream survival prediction tasks.

### E Computing Resources

All experiments were conducted on a high-performance computing cluster running AlmaLinux. For the largest language models (DSL33 and L33), training required three NVIDIA A100 GPUs (80 GB each; total 240 GB GPU memory), while inference was performed on two A100 GPUs. The encoder models were trained and evaluated on a single NVIDIA A100 GPU (80 GB). After getting the embedding vectors from LLMs, survival models were trained and evaluated on a workstation equipped with an NVIDIA RTX 4090 GPU. System memory usage was 32 GB RAM. Software dependencies, including specific versions of Python libraries and frameworks, are documented in `environment_tts_forecasting.yml` file of our *anonymized* repository in the *Supplementary Material*.

### F LLM prompt to generate clinical textual time-series from PMOA case reports

#### Prompt

You are a physician.

Extract the clinical events and the related time stamp from the case report. The admission event has timestamp 0. If the event is not available, we treat the event, e.g. current main clinical diagnosis or treatment with timestamp 0. The events that happened before event with 0 timestamp have negative time, the ones after the event with 0 timestamp have positive time. The timestamp are in hours. The unit will be omitted when output the result. If there is no temporal information of the event, please use your knowledge and events with temporal expression before and after the events to provide an approximation. We want to predict the future events given the events happened in history. For example, here is the case report.

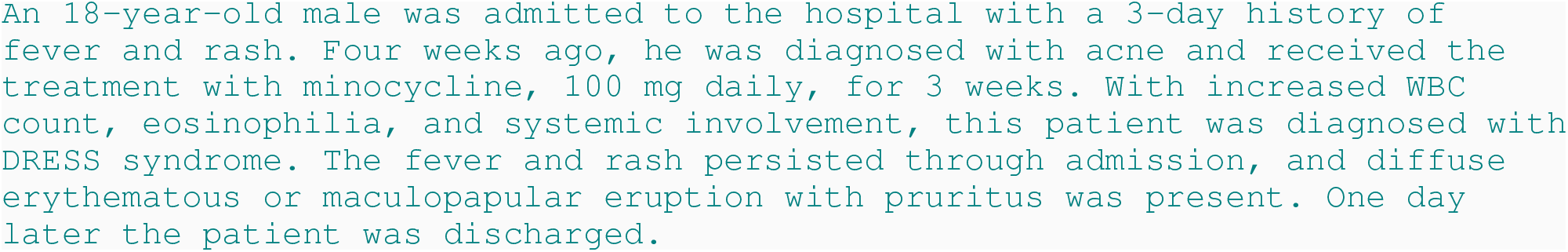

Let’s find the locations of event in the case report, it shows that four weeks ago of fever and rash, four weeks ago, he was diagnosed with acne and receive treatment. So the event of fever and rash happened four weeks ago, 672 hours, it is before admitted to the hospital, so the time stamp is -672. diffuse erythematous or maculopapular eruption with pruritus was documented on the admission exam, so the timestamp is 0 hours, since it happens right at admission. DRESS syndrome has no specific time, but it should happen soon after admission to the hospital, so we use our clinical judgment to give the diagnosis of DRESS syndrome the timestamp 0. then the output should look like:

~~~
18 years old | 0 male | 0
admitted to the hospital | 0
fever | -72
rash | -72
acne | -672
minocycline | -672
increased WBC count | 0
eosinophilia | 0
systemic involvement | 0
diffuse erythematous or maculopapular eruption | 0
pruritis | 0
DRESS syndrome | 0
fever persisted | 0
rash persisted | 0
discharged | 24
~~~

Separate conjunctive phrases into its component events and assign them the same timestamp (for example, separation of ‘fever and rash’ into 2 events: ‘fever’ and ‘rash’). If the event has duration, assign the event time as the start of the time interval. Attempt to use the text span without modifications except ‘history of’ where applicable. Include all patient events, even if they appear in the discussion; do not omit any events; include termination/discontinuation events; include the pertinent negative findings, like ‘no shortness of breath’ and ‘denies chest pain’. Show the events and timestamps in rows, each row has two columns: one column for the event, the other column for the timestamp. The time is a numeric value in hour unit. The two columns are separated by a pipe | as a bar-separated file. Skip the title of the table. Reply with the table only.

### G Prompting strategies for forecasting tasks

#### F.1 Zero-shot

##### Prompt

You are an expert physician.

Reply to the prompt with structured predictions in a k-item, bar-separated row.

For example, if there k=3 events (A, B, C) and only B occurs in the time window, then: 0 | 1 | 0 would be the correct response.

Reply with the answer only. Do NOT provide any other text. Do NOT explain.

#### F.2 Zero-shot (with degree of confidence)

##### Prompt

You are an expert physician.

Reply to the prompt with structured predictions in a k-item, bar-separated row. Assess whether the event happens in the time window using a scale from 1 to 5, where 5 means it definitely happened and 1 means it did not happen. response.

Reply with the answer only. Do NOT provide any other text. Do NOT explain.

For example, if there k=3 events (A, B, C) and only B occurs in the time window, then: 1|5|1 would be the correct

#### F.3 Few-shot

##### Prompt

You are an expert physician.

Reply to the prompt with predictions in a k-item, bar-separated row to answer if the k events happen within a given forecast time window.

Example 1. If there k=3 events (A, B, C) and only B occurs in the requested time window, then: 0 | 1| 0 would be the correct response.

Example 2. Suppose the time series is:

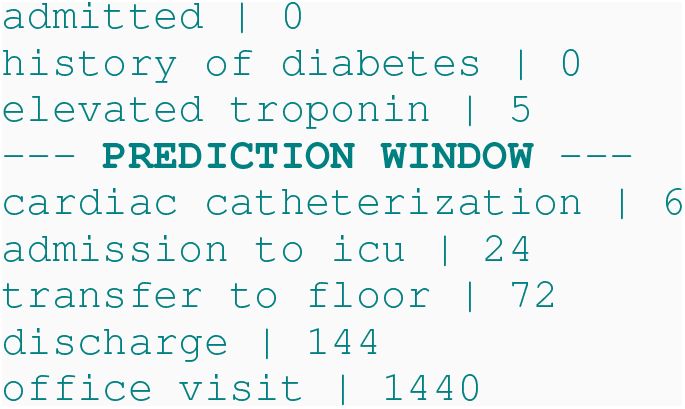

Then, if the prediction is at 5 hours, the time preceding “— PREDICTION WINDOW —”, the events admitted, history of diabetes, and elevated troponin have occurred. Supposing k=4 and a forecast window of 24 hours, then the task it to determine which of cardiac catheterization, admission to icu, transfer to floor, and discharge happen. So the correct response for the example is: 1 |1| 0 | 0 because cardiac catherization (6) and admission to icu (24) happen before 5 + 24 = 29.

Reply with the answer only. Do NOT provide any other text. Do NOT explain.

#### F.4 Few-shot ordinal

##### Prompt

You are an expert physician.

Reply to the prompt with predictions in a k-item, bar-separated row to answer if the k events happen within a given forecast time window, using a scale from 1 to 5, where 5 means it definitely happened and 1 means it did not happen.

Example 1. If there k=3 events (A, B, C) and only be the correct response.

Example 2. Suppose the time series is:

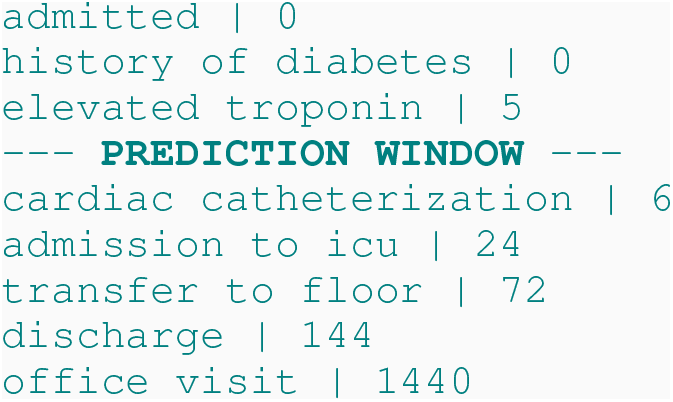

Then, if the prediction is at 5 hours, the time preceding “— PREDICTION WINDOW —”, the events admitted, history of diabetes, and elevated troponin have occurred. Supposing k=4 and a forecast window of 24 hours, then the task it to determine which of cardiac catheterization, admission to icu, transfer to floor and discharge happen. So the an excellent response for the example B occurs in the time window, then: 1 | 5 | 1 would is: 5 | 4 | 1 | 1 because cardiac catherization (6) and admission to icu (24) happen before 5 + 24 = 29.

Reply with the answer only. Do NOT provide any other text. Do NOT explain.

### H Open-source models for avoiding potential data leakage

We recognize that data leakage is a critical concern in evaluating model performance, particularly when using openaccess text corpora. Below, we outline key factors that mitigate this concern in our study:

1. **Temporal annotations—not raw text—drive forecasting tasks**. The core contribution of this work lies in the temporal structuring of clinical narratives. While large language models (LLMs) such as *DeepSeek* and *Llama-3*.*3* may have encountered PubMed Open Access (PMOA) case reports during pretraining, they were never exposed to the extracted timestamps or temporally ordered and future-masked (event, time) tuples used in our evaluations. These derived temporal annotations are essential to all forecasting tasks in our study—including F1-based next-event prediction, temporal ordering, and survival analysis—and constitute novel structured data not present in the original text.
2. **Temporal reordering fundamentally alters the learning problem**. Even if LLMs were pretrained on PMOA narratives, they would have encountered events in their original, text-based order. In contrast, our framework chronologically reorders events based on extracted timestamps, creating a new temporal structure. This transformation introduces a distinct modeling challenge, as it breaks the sequential narrative flow seen during pretraining. As shown in Section 4.4.1, training on time-ordered versus text-ordered annotations leads to measurable differences in performance, underscoring the impact of this restructuring.
3. **External validation reveals limited effects of memorization**. Our experimental design separates internal (T2S2 test set) and external validation (sepsis-10 and sepsis-100 cohorts), using entirely held-out case reports. The performance drop observed on external datasets (Tables 1–2) suggests that any potential memorization of PMOA case reports during pretraining yields minimal benefit and reinforces the robustness of our empirical findings.
4. **Broader context: standard practices and safeguards**. Training data overlap is a general challenge in modern machine learning, especially for LLMs trained on web-scale corpora. Our work adheres to community standards while incorporating additional safeguards—such as explicit temporal annotation, event reordering, and external dataset validation—that reduce the risk of leakage and differentiate our tasks from standard text modeling. To address concerns regarding potential pretraining overlap, we evaluated multiple models with documented training corpora. Both *OLMO-32B-Instruct* (Allen Institute for AI) and *RedPajama-INCITE-7B-Instruct* represent fully opensource models with transparent training data. Notably, RedPajama explicitly excludes PubMed from its training corpus, making it an ideal benchmark for assessing causal leakage. The comparable performance of these models to those potentially exposed to PMOA (see point 5 above) provides strong evidence that our temporal forecasting tasks are meaningfully distinct from standard language modeling on clinical narratives.
5. **Empirical evidence from PubMed-excluded models**. Our results provide direct empirical evidence against meaningful causal leakage. *RedPajama-INCITE-7B-Instruct*, which explicitly excludes PubMed from its training corpus, achieves comparable performance to other decoder models (0.352 F1 at 24h, 0.618 concordance) that potentially had access to PMOA during pretraining. This similarity in performance across models with and without PubMed exposure demonstrates that our T2S2 prediction task—with its temporal restructuring, event masking, and partial history inputs—is sufficiently distinct from raw case report text that prior exposure provides no meaningful advantage. Furthermore, *OLMO-32B-Instruct* shows competitive performance (0.411 F1 at 24h), providing additional validation from a model with fully documented training data.

### I Broader Impact

This work advances equitable, scalable, and transparent medical AI by introducing a framework for forecasting clinical risk in sepsis from narrative documentation. Sepsis remains a leading cause of preventable mortality worldwide, with outcomes highly dependent on timely recognition and intervention. In many under-resourced healthcare settings, structured EHR data and real-time specialist input are unavailable, leaving unstructured narratives as the primary record of patient care. Our approach transforms such narratives into temporally structured representations, enabling systematic evaluation of forecasting methods for short- and long-horizon event prediction, event ordering, and survival analysis. By releasing the dataset, extraction pipeline, and evaluation framework, we lower barriers for developing and benchmarking temporal forecasting models in sepsis, and promote reproducibility in LLM-based clinical information extraction.

However, societal considerations are critical. LLM-assisted extraction can introduce inaccuracies or hallucinations, and unvalidated forecasts risk amplifying bias or error. Although timelines are derived from non-identifiable public case reports, their interpretation could still reinforce historical biases if not carefully contextualized. Forecasting results from case reports may also differ from those using routine clinical notes, limiting generalizability. Moreover, applying these forecasting models in high-stakes clinical decision-making without expert oversight poses safety risks. Responsible use requires transparent communication of dataset and model limitations, explicit uncertainty reporting, and safeguards to prevent inappropriate deployment in patient-facing settings.

## Footnotes

1 Our code is available at https://github.com/Shahriarnz14/Textual-Time-Series-Forecasting

